# Detection of COVID-19 Infection from Routine Blood Exams with Machine Learning: a Feasibility Study

**DOI:** 10.1101/2020.04.22.20075143

**Authors:** Davide Brinati, Andrea Campagner, Davide Ferrari, Massimo Locatelli, Giuseppe Banfi, Federico Cabitza

## Abstract

**Background:** The COVID-19 pandemia due to the SARS-CoV-2 coronavirus, in its first 4 months since its outbreak, has to date reached more than 200 countries worldwide with more than 2 million confirmed cases (probably a much higher number of infected), and almost 200,000 deaths. Amplification of viral RNA by (real time) reverse transcription polymerase chain reaction (rRT-PCR) is the current gold standard test for confirmation of infection, although it presents known shortcomings: long turnaround times (3-4 hours to generate results), potential shortage of reagents, false-negative rates as large as 15-20%, the need for certified laboratories, expensive equipment and trained personnel. Thus there is a need for alternative, faster, less expensive and more accessible tests.

**Material and methods:** We developed two machine learning classification models using hematochemical values from routine blood exams (namely: white blood cells counts, and the platelets, CRP, AST, ALT, GGT, ALP, LDH plasma levels) drawn from 279 patients who, after being admitted to the San Raffaele Hospital (Milan, Italy) emergency-room with COVID-19 symptoms, were screened with the rRT-PCR test performed on respiratory tract specimens. Of these patients, 177 resulted positive, whereas 102 received a negative response.

**Results:** We have developed two machine learning models, to discriminate between patients who are either positive or negative to the SARS-CoV-2: their accuracy ranges between 82% and 86%, and sensitivity between 92% e 95%, so comparably well with respect to the gold standard. We also developed an interpretable Decision Tree model as a simple decision aid for clinician interpreting blood tests (even off-line) for COVID-19 suspect cases.

**Discussion:** This study demonstrated the feasibility and clinical soundness of using blood tests analysis and machine learning as an alternative to rRT-PCR for identifying COVID-19 positive patients. This is especially useful in those countries, like developing ones, suffering from shortages of rRT-PCR reagents and specialized laboratories. We made available a Web-based tool for clinical reference and evaluation^1^.

## 1 Introduction

The pandemic disease caused by the SARS-CoV-2 virus named COVID-19 is requiring unprecedented responses of exceptional intensity and scope to more than 200 states around the world, after having infected, in the first 4 months since its outbreak, a number of people between 2 and 20 million with at least 200,000 deaths. To cope with the spread of the COVID-19 infection, governments all over the world has taken drastic measures like the quarantine of hundreds of millions of residents worldwide.

However, because of the COVID-19 symptomatology, which showed a large number of asymptomatics [12], these efforts are limited by the problem of differentiating between COVID-19 positive and negative individuals. Thus, tests to identify the SARS–CoV-2 virus are believed to be crucial to identify positive cases to this infection and thus curb the pandemic.

To this aim, the current test of choice is the reverse transcriptase Polymerase Chain Reaction (rt-PCR)-based assays performed in the laboratory on respiratory specimens. Taking this as a gold standard, machine learning techniques have been employed to detect COVID-19 from lung CT-scans with 90% sensitivity, and high AUROC (0.95) [25, 18]. Although chest CTs have been found associated with high sensitivity for the diagnosis of COVID-19 [1], this kind of exam can hardly be employed for screening tasks, for the radiation doses, the relative low number of devices available, and the related operation costs. A similar attempt was recently performed on chest x-rays [4], which is a low-dose and less expensive test, with promising statistical performance (e.g., sensitivity 97%). However, since almost 60% of chest x-rays taken in patients with confirmed and symptomatic COVID-19 have been found to be normal [40], systems based on this exam need to be thoroughly validated in real-world settings [6].

The public health emergency requires an unprecedented global effort to increase testing capacity [29]. The large demand for rRT-PCR tests (also commonly known as nasopharyngeal swab tests) due to the worldwide extension of the virus is highlighting the limitations of this type of diagnosis on a large-scale such as: the long turnaround times (on average over 2 to 3 hours to generate results); the need of certified laboratories; trained personnel; expensive equipment and reagents for which demand can easily overcome supply [26]. For instance in Italy, the scarcity of reagents and specialized laboratories forced the government to limit the swab testing to those people who clearly showed symptoms of severe respiratory syndrome, thus leading to a number of infected people and a contagion rate that were largely underestimated [34].

For this reason, and also in light of the predictable wide adoption of mobile apps for contact tracing [14], which will likely increase the demand for population screening, there is an urgent need for alternative (or complementary) testing methods by which to quickly identify infected COVID-19 patients to mitigate virus transmission and guarantee a prompt patients treatment.

On a previous work published in the laboratory medicine literature [13], we showed how simple blood tests might help identifying false positive/negative rRT-PCR tests. This work and the considerations made above strongly motivated us to apply machine learning methods to routine, low-cost^2^ blood exams, and to evaluate the feasibility of predictive models in this important task for the mass-screening of potential COVID-19 infected individuals. In what follows we report this feasibility study in detail.

## 2 Methods

The aim of this work is to develop a predictive model, based on Machine Learning techniques, to predict the positivity or negativity for COVID-19. In the rest of this Section we report on the dataset used for model training and on the data analysis pipeline adopted.

### 2.1 Data Description

The dataset used for this study was made available by the *IRCCS Ospedale San Raffaele*^3^ and it consisted of 279 cases, randomly extracted from patients admitted to that hospital from the end of February 2020 to mid of March 2020. Each case included the patient’s age, gender, and values from routine blood tests, as well as the result of the RT-PCR test for COVID-19, performed by nasopharyngeal swab. The parameters collected by the blood test are reported in Table 1.

**Table 1.** Features of the dataset considered in the present study.

| Feature | Data Type |
| --- | --- |
| Gender | Categorical |
| Age | Numerical (discrete) |
| Leukocytes (WBC) | Numerical (continuous) |
| Platelets | Numerical (continuous) |
| C-reactive Protein (CRP) | Numerical (continuous) |
| Transaminases (AST) | Numerical (continuous) |
| Transaminases (ALT) | Numerical (continuous) |
| Gamma Glutamyl Transferasi (GGT) | Numerical (continuous) |
| Lactate dehydrogenase (LDH) | Numerical (continuous) |
| Neutrophils | Numerical (continuous) |
| Lymphocytes | Numerical (continuous) |
| Monocytes | Numerical (continuous) |
| Eosinophils | Numerical (continuous) |
| Basophils | Numerical (continuous) |
| Swab | Categorical |

<sup>2</sup> A qualitative estimation of the cost of the exams used for this study is 15 euros per test, approximately five times cheaper than rt-PCR testing.
<sup>3</sup> IRCCS is the Italian acronym for Scientific Institute for Research, Hospitalization and Healthcare

The dependent variable “*Swab*” is binary and it is equal to 0 in the absence of COVID-19 infection (negative swab test), and it is equal to 1 in the case of COVID-19 infection (positive to the swab test). The number of occurrences for the negative and positive class was respectively 102 (37%) and 177 (63%), thus the dataset was slightly imbalanced towards positive cases.

Figure 1 shows the pairwise correlation of the features used for this study, while Figure 2 focuses on variables “*Age*”, “*WBC* “, “*CRP* “, “*AST*” and “*Lymphocytes*”.

**Fig. 1.**
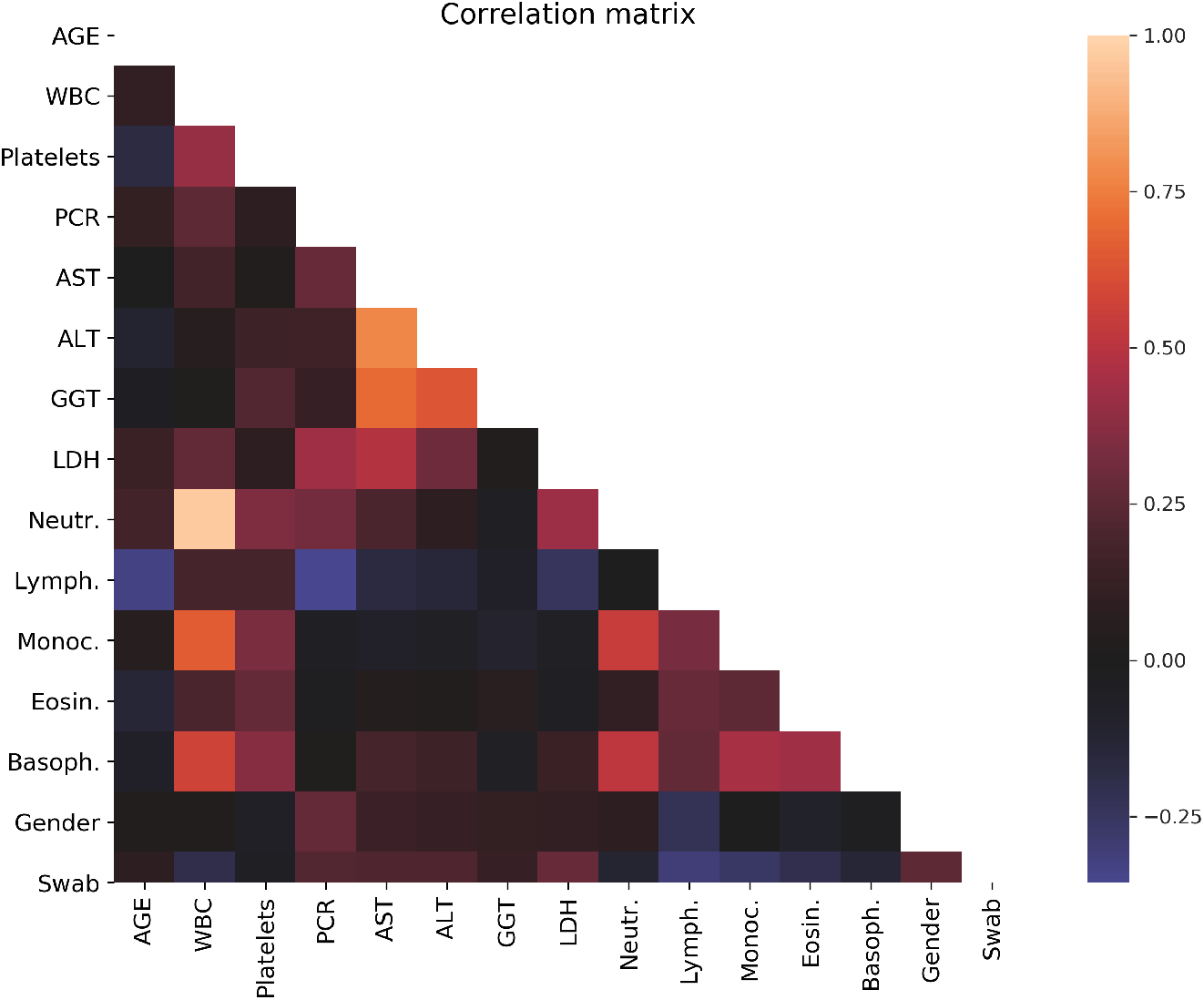
Pairwise Pearson correlation of the features taken into account for this case study.

**Fig. 2.**
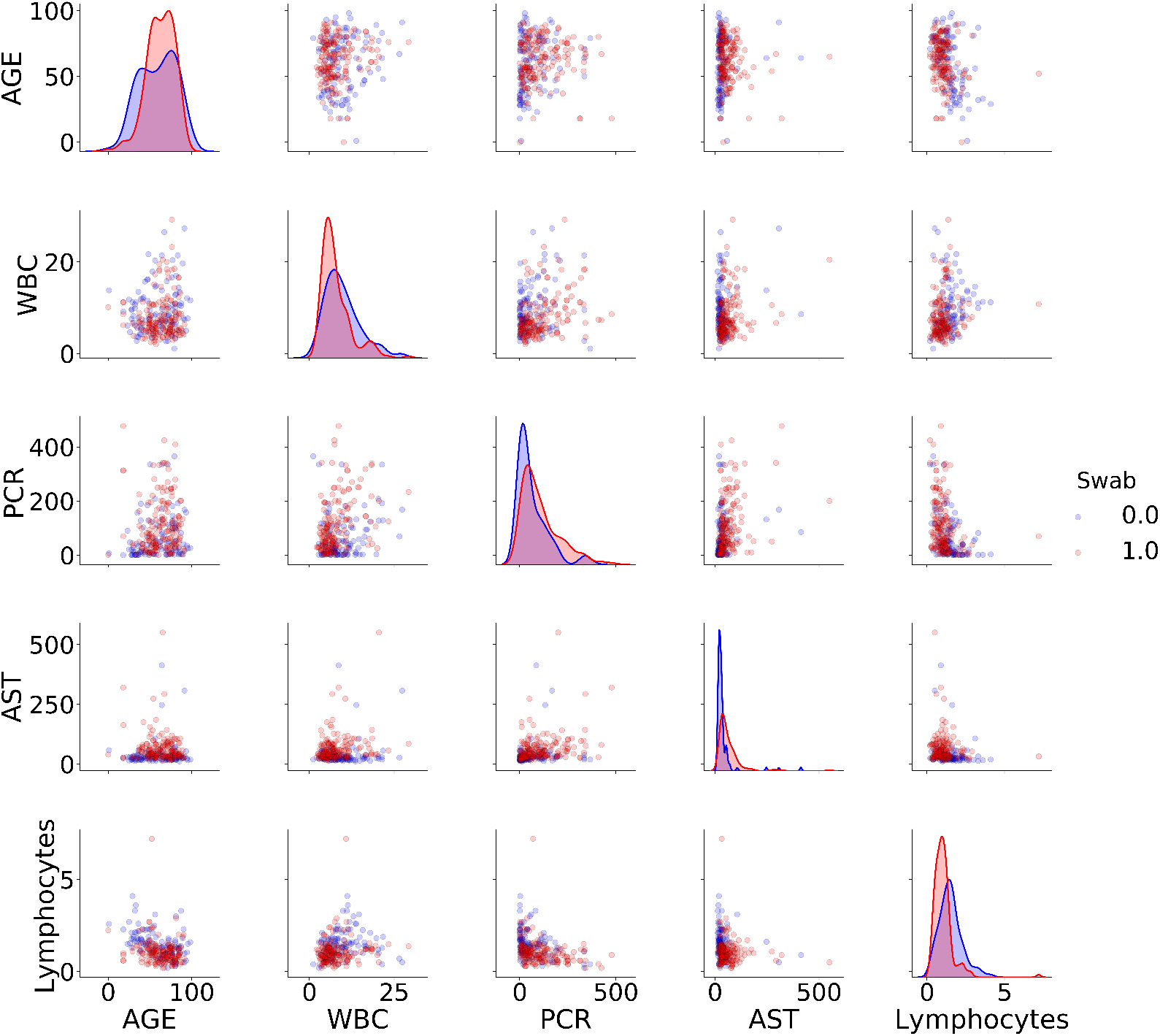
Distribution plots and pairwise scatter plots of selected features. Red points and red distributions represent positive patients to Covid19, while blue points represent negative patients.

### 2.2 Data Manipulation

First of all, the categorical feature *Gender* has been transformed into two binary features by *one-hot encoding*. Further, we notice that the dataset was affected by missing values in most of its features (see Table 2).

**Table 2.** Features and missing values in the dataset

| Feature | N° of missing | % of missing on the total |
| --- | --- | --- |
| C-reactive protein (CRP) | 6 | 2.1 |
| Aspartate Aminotransferase (AST) | 2 | 0.7 |
| Alanine Amino Transferase (ALT) | 13 | 4.6 |
| Gamma Glutamyl Transferase (GGT) | 143 | 51.2 |
| Lactate Dehydrogenase (LDH) | 85 | 30.4 |
| Leukocyte Count (WBC) | 2 | 0.7 |
| Platelets | 2 | 0.7 |
| Neutrophils | 70 | 25 |
| Lymphocytes | 70 | 25 |
| Monocytes | 70 | 25 |
| Eosinophils | 70 | 25 |
| Basophils | 71 | 25.4 |

To address data incompleteness, we performed missing data imputation by means of the *Multivariate Imputation by Chained Equation* (MICE) [5] method. MICE is a multiple imputation method that works in an iterative fashion: in each imputation round, one feature with missing values is selected and is modeled as a function of all the other features; the estimated values are then used to impute the missing values and re-used in the subsequent imputation rounds.

We chose this method because multiple imputation techniques are known to be more robust and better capable to account for uncertainty compared with single imputation ones [33] (as they employ the joint distribution of the available features), and MICE in particular can also handle different data types.

### 2.3 Model Training, Selection and Evaluation

We developed and compared different classes of Machine Learning classifiers. In particular, we considered the following classifier models:

– *Decision Tree* [35] *(DT);*
– *Extremely Randomized Trees* [16] *(ET);*
– *K-nearest neighbors* [2] *(KNN);*
– *Logistic Regression* [20] *(LR);*
– *Naïve Bayes* [23] *(NB);*
– *Random Forest* [21] *(RF);*
– *Support Vector Machines* [36] (SVM).

We also considered a modification of the Random Forest algorithm, called three-way Random Forest classifier [7] (TWRF), which allows the model to abstain on instances for which it can express low confidence; in so doing, a TWFR achieves higher accuracy on the effectively classified instances at expense of coverage (i.e., the number of instances on which it makes a prediction). We decided to consider also this class of models as they could provide more reliable predictions in a large part of cases, while exposing the uncertainty regarding other cases so as to suggest further (and more expensive) tests on them.

From a technical point of view, since Random Forest is a class of probability scoring classifiers (that is, for each instance the model assigns a probability score for every possible class), the abstention is performed on the basis of two thresholds *α, β* ∈ [0, 1]: if we denote with 1 the positive class and 0 the negative class, then each instance is classified as positive if *score*(1) *> α* and *score*(1) *> score*(0), negative if *score*(0) *> β* and *score*(0) *> score*(1) and, otherwise, the model abstains. In these models the performance is usually evaluated only on the non-abstained instances [15], and the coverage is a further performance element to be considered. The models mentioned above have been trained, and evaluated, through a *nested cross validation* [19, 9] procedure. This procedure allows for an unbiased generalization error estimation while the hyperparameter search (including feature selection) is performed: an inner cross-validation loop is executed to find the optimal hyperparameters via grid search and an outer loop evaluates the model performance on five folds.

Models were evaluated in terms of *accuracy, balanced accuracy* ^4^, *Positive Predictive Value* (PPV)^5^, *sensitivity, specificity* and, except for the three-way Random Forest, the *area under the ROC curve* (AUC). After discussing this with the clinicians involved in this study, we considered accuracy and sensitivity to be the main quality metrics, since false negatives (that is, patients positive to COVID-10 which are, however, classified as negative, and possibly let go home) are more harmful than false positives in this screening task.

## 3 Results

Tables 3 and 4 show the 95% confidence intervals of, respectively, average accuracy and average balanced accuracy (that is, the average of sensitivity and specificity) of the models (on the nested cross-validation) trained on the two best-performing sets of features: the first one, dataset A, includes all the variables, while the second one, dataset B, excludes the “*Gender*” variable, as this was found of negligible predictive value.

**Table 3.** The models’ performance: 95% C.I. of model accuracy on *5-folds nested CV*.

|  | DT | ET | KNN | LR | NB | RF | SVM | TWRF |
| --- | --- | --- | --- | --- | --- | --- | --- | --- |
| A (all features) | [0.70, 0.78] | [0.68, 0.79] | [0.66, 0.76] | [0.70, 0.81] | [0.64, 0.81] | [0.74, 0.80] | [0.69, 0.80] | **[0.83, 0.89]** |
| B (without Gender) | [0.62, 0.75] | [0.74, 0.82] | [0.66, 0.76] | [0.670, 0.79] | [0.65, 0.76] | **[0.71, 0.86]** | [0.66, 0.79] | **[0.83, 0.89]** |

**Table 4.** The models’ performance: 95% C.I. of model balanced accuracy on *5-folds nested CV*.

|  | DT | ET | KNN | LR | NB | RF | SVM | TWRF |
| --- | --- | --- | --- | --- | --- | --- | --- | --- |
| A (all features) | [0.64, 0.71] | [0.67, 0.81] | [0.60, 0.74] | [0.65, 0.79] | [0.63, 0.77] | [0.70, 0.82] | [0.69, 0.76] | **[0.83, 0.87]** |
| B (without Gender) | [0.63, 0.73] | [0.67, 0.84] | [0.61, 0.74] | [0.64, 0.74] | [0.63, 0.76] | **[0.70, 0.80]** | [0.65, 0.77] | **[0.83, 0.87]** |

Figure 3 shows the performance of the traditional models (i.e., the TWRF model was excluded) on the *nested cross-validation*.

**Fig. 3.**
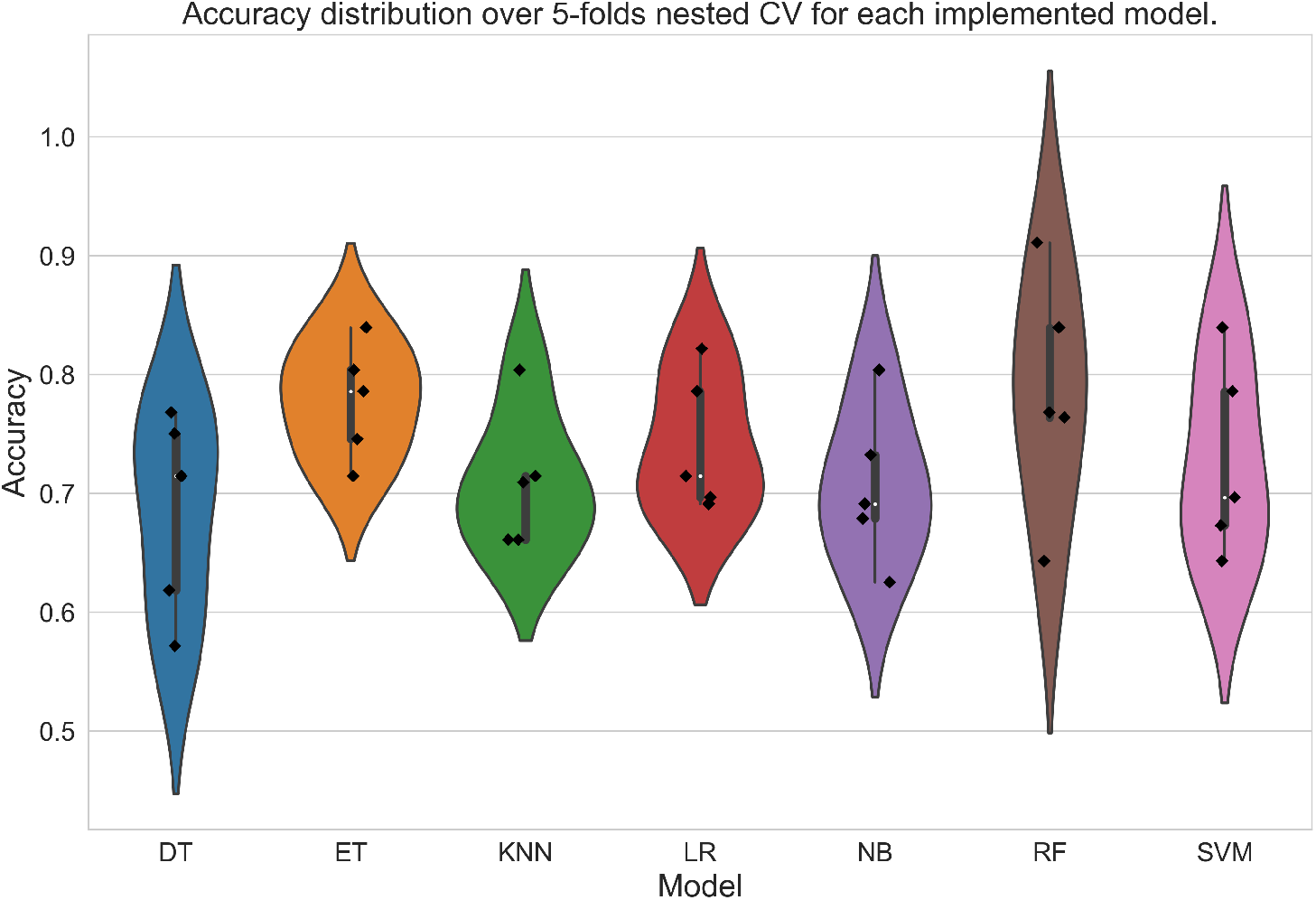
Violin plots of the accuracy distributions reached by each models on five folds (on dataset B).

To further validate the above findings, the entire dataset has been splitted into training and test/validation sets, respectively the 80% and the 20% of the total instances. The best performing model, i.e. the Random Forest classifier, trained on dataset B, achieved the following results on the test/validation set: accuracy = 82%, sensitivity = 92%, PPV = 83%, specificity = 65%, AUC = 84%. Figures 4 and 5 show the performance of this model in the ROC and precision/recall space, respectively.

**Fig. 4.**
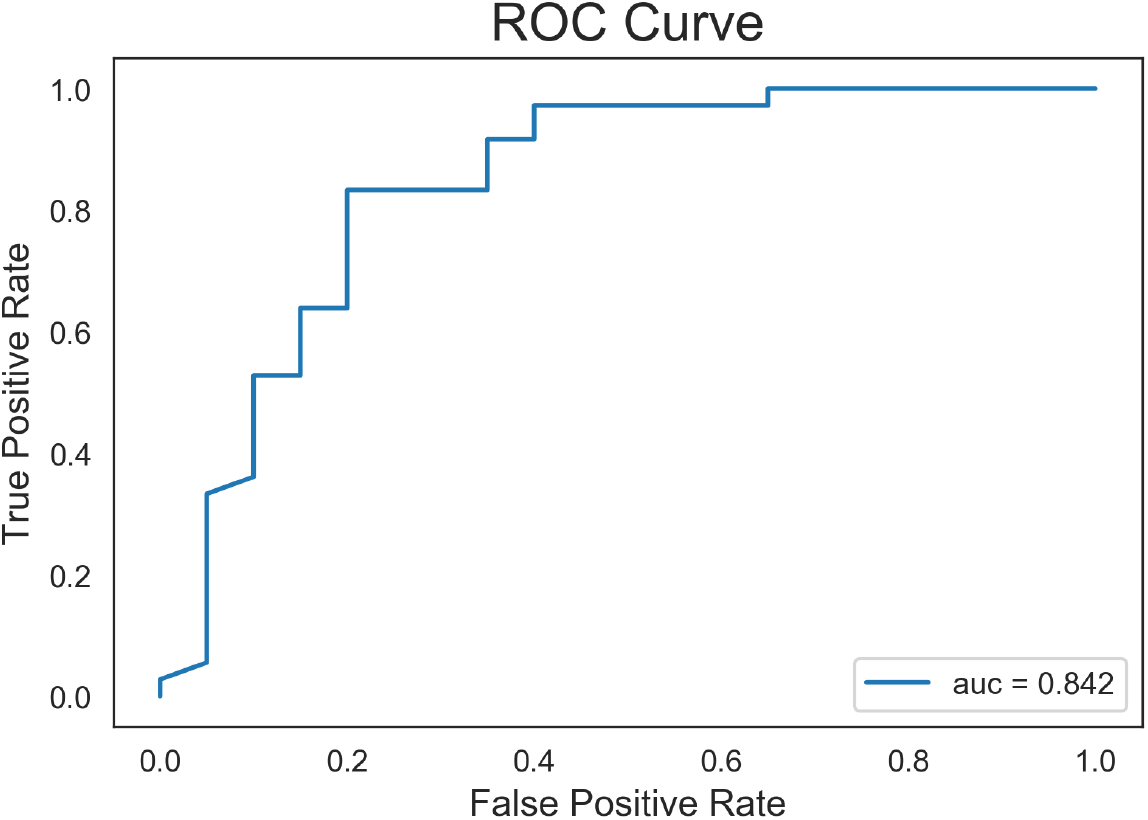
The sensitivity and specificity curve (i.e., sensitivity /positive predictive value curve or, equivalently true positive rate / false positive rate as depicted in the Figure) and the area under the ROC curve.

**Fig. 5.**
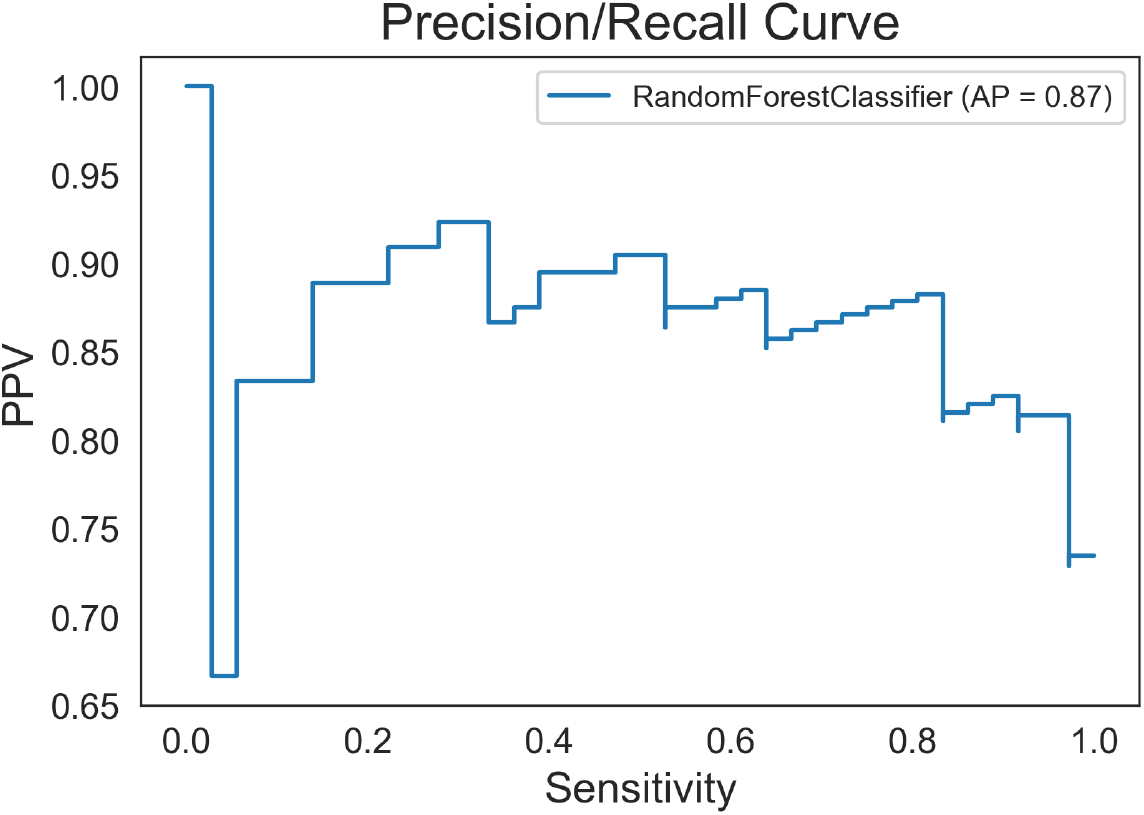
The precision/recall (i.e., positive predictive value / sensitivity curve) and its area.

The optimal hyperparameters found are shown in Table 5.

**Table 5.** Optimal hyperparameters for the Random Forest classifier. For the sake of reproducibility, also the random seed is reported.

| Hyperparameters | Value |
| --- | --- |
| Max Depth | - |
| Criterion | Gini |
| $N^\circ$ estimators | 100 |
| Random seed for reproducibility | 123 |

Similarly, for the best three-way Random Forest classifier on the validation set we observed: accuracy = 86%, sensitivity = 95%, PPV = 86%, specificity = 75%, coverage = 70% (that is, for 30% of the validation instances the model abstained). The feature importance assessed for the the best performing model (Random Forest on dataset B), are shown in Figure 6.

**Fig. 6.**
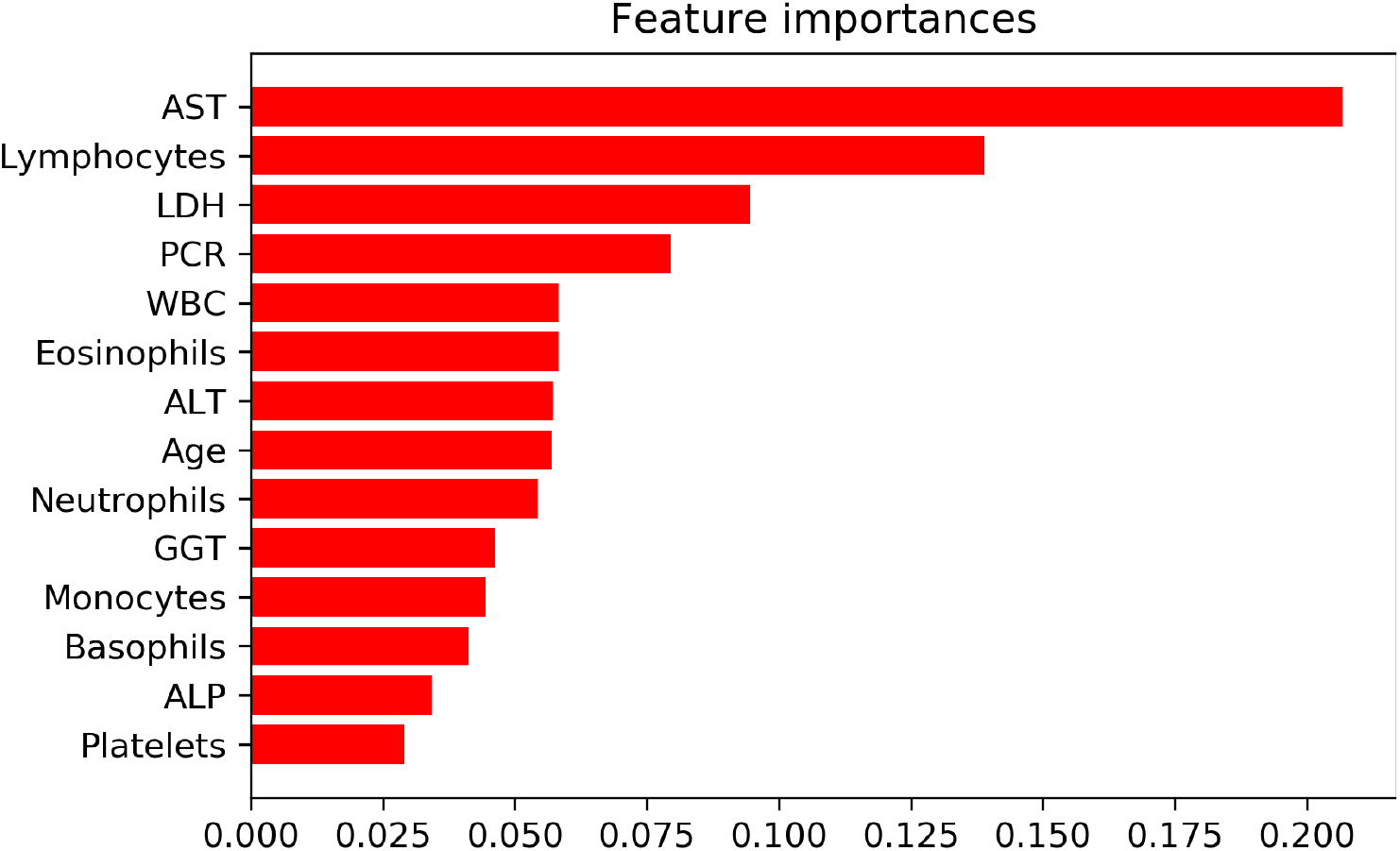
Feature importance scores for the best performing model.

In order to provide an interpretable overview (in the sense of eXplainable AI[17]) of the predictive models that we developed, we also developed a Decision Tree model, which is shown in Figure 7. Although the depicted decision tree is associated with a lower discriminative performance than the two former (inscrutable) models, such a tree can be used as a simple decision aid by clinicians interested in the use of blood values to assess COVID-19 suspect cases.

**Fig. 7.**
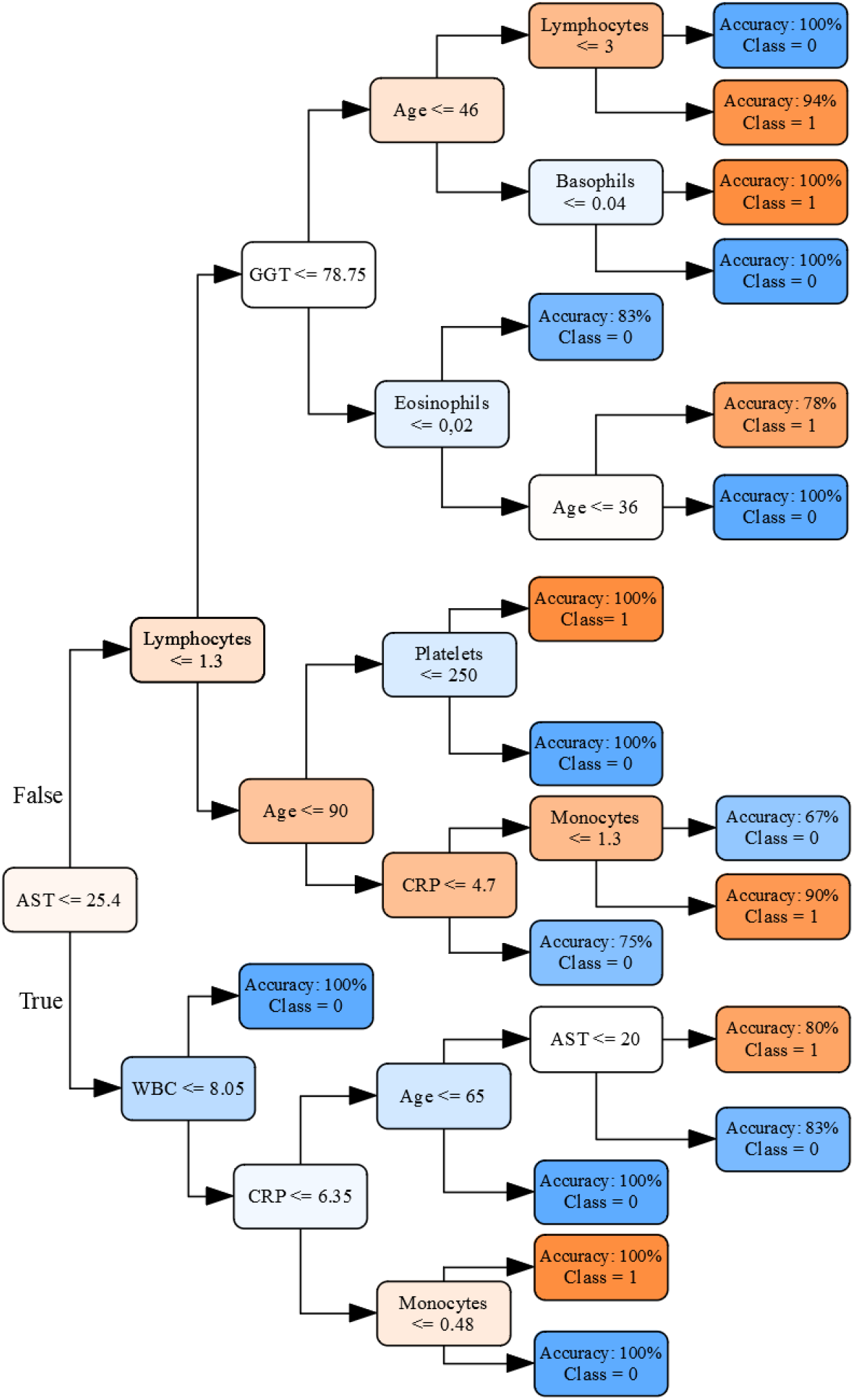
An interpretable Decision Tree, developed in order to support the interpretation of the predictions from the other models. Color gradients denote predictivity for either classes (shades of blue correspond to COVID-19 negativity, shades of orange to positivity).

## 4 Discussion

We have developed two machine learning models to discriminate between patients who are either positive or negative to the SARS-CoV-2, which is the coronavirus causing the COVID-19 pandemia. In this task, patients are represented in terms of few basic demographic characteristics (gender, age) and a small array of routine blood tests, chosen for their convenience, low cost and because they are usually available within 30 minutes from the blood draw in regular emergency department. The ground truth was established through RT-PCR swab tests.

We presented the best traditional model, as it is common practice, and a three-way model, which guarantees best sensitivity and positive predictive value: the former is the proportion of infected (and contagious) people who will have a positive result and therefore it is useful to clinicians when deciding which test to use. On the other hand, PPV is useful for patients as it tells the odds of one having COVID-19 if they have a positive result.

The performance achieved by these two best models (sensitivity between 92% and 95%, accuracy between 82% and 86%) provides proof that this kind of data, and computational models, *can be used* to discriminate among potential COVID-19 infectious patients with sufficient reliability, and similar sensitivity to the current Gold Standard. This is the most important contribution of our study.

Also from the clinical point of view, the feature selection was considered valid by the clinicians involved. Indeed, the specialist literature has found that COVID-19 positivity is associated with lymphopenia (that is, abnormally low level of white blood cells in the blood), damage to liver and muscle tissue [42, 39], and significantly increased C-reactive protein (CRP) levels [10]. In [27] a comprehensive list of the most frequent abnormalities in COVID-19 patients has been reported: among the 14 conditions considered, they report increased aspartate aminotransferase (AST), decreased lymphocyte count (WBC), increased lactate dehydrogenase (LDH), increased C-reactive protein (CRP), increased white blood cell count (WBC) and increased alanine aminotransferase (ALT).

These parameters are also the most predictive features identified by the best classifier (Random Forest), all together with the Age attribute. Also other studies confirm the relevance of these features and their association with the COVID-19 positivity [8, 30, 32, 44], compared to other kinds of pneumonia [43]. This also gives confirmation that our models ground on clinically relevant features and that most of these values can be extracted from routine blood exams.

The interpretable Decision Tree model provides a further confirmation (see Figure 7) of the soundness of the approach: the clinicians (ML, GB) and the biochemist (DF) involved in this study found reasonable that the AST would be the first parameter to consider (i.e., mirrored by the fact that AST was the root of the decision tree) and that it was found to be the most important predictive feature. Indeed, values of AST below 25 are good predictors of COVID-19 positivity (accuracy = PPV = 76%), while values below 25 are a good predictor of COVID-19 negativity (accuracy = Negative Predictive Value = 83%). Similar observations can also be made about CRP, Lymphocytes and general WBC counts.

No statistically significant difference was found between the accuracy and the balanced accuracy of the models (as mirrored by the overlap of the 95% confidence intervals), as a sign that the dataset was not significantly unbalanced.

Moreover, we can notice that the best performing ML classifier (Random Forest) exhibited a very high sensitivity (*∼* 90%) but, in comparison, a limited specificity of only 65%. That gives the main motivation for the three-way classifier: this model offers a trade-off between increased specificity (a 10% increment compared with the best traditional ML model) and reduced coverage, as the three-way approach abstains on uncertain instances (i.e., the cases that cannot be classified with high confidence neither as positive nor negative). This means that the model yields more robust and reliable prediction for the classified instances (as it is mirrored by the increase in all of the performance measures), while for the other ones it is anyway useful in suggesting further tests, e.g., by either a PCR-RNA swab test or a chest x-ray.

In regard to the specificity exhibited by our models, we can further notice that even while these values are relatively low compared with other tests (which are more specific but slower and less accessible), this may not be too much of a limitation as there is a significant disparity between the costs of false positives and false negatives and in fact our models favors sensitivity (thus, they avoid false negatives). Further, the high PPV (*>* 80%) of our models suggest that the large majority of cases identified as positives by our models would likely be COVID-19 positive cases.

That said, the study presents two main limitations: the first, and more obvious one, regards the relatively low number of cases considered. This was tackled by performing nested cross-validation in order to control for bias [38], and by employing models that are known to be effective also with moderately sized samples [3, 31, 37]. Nonetheless, further research should be aimed at confirming our findings, by integrating hematochemical data from multiple centers and increasing the number of the cases considered. The second limitation may be less obvious, as it regards the reliability of the ground truth itself. Although this was built by means of the current gold standard for COVID-19 detection, i.e., the rRt-PCR test, a recent study observed that the accuracy of this test may be highly affected by problems like inadequate procedures for collection, handling, transport and storage of the swabs, sample contamination, and presence of interfering substances, among the others [28]. As a result, some recent studies have reported up to 20% false-negative results for the rRt-PCR test [41, 24, 22], and a recent systematic review reported an average sensitivity of 92% and cautioned that “up to 29% of patients could have an initial RT-PCR false-negative result”. Thus, contrary to common belief and some preliminary study (e.g., [11]), the accuracy of this test could be less than optimal, and this could have affected the reliability of the ground truth also in this study (as in any other using this test for ground truthing, unless cases are annotated after multiple tests. However, besides being a limitation, this is also a further motivation to pursue alternative ways to perform the diagnosis of SARS-CoV-2 infection, such as our methods are.

Future work will be devoted to the inclusion of more hematochemical parameters, including those from arterial blood gas assays (ABG), to evaluate their predictiveness with respect to COVID-19 positiveness, and the inclusion of cases whose probability to be COVID-positive is almost 100%, as they resulted positive to two or more swabs or to serologic antibody tests. This would allow to associate a higher weight with misidentifying those cases, so as, we conjecture, improve the sensitivity further.

Moreover, we want to investigate the interpretability of our models further, by both having more clinicians validate the current Decision Tree, and possibly construct a more accurate one, so that clinicians can use it as a convenient decision aid to interpret blood tests in regard to COVID-19 suspect cases (even off-line).

Finally, this was conceived as a feasibility study for an alternative COVID-19 test on the basis of hematochimical values. IN virtue of this ambitious goal, the success of this study does not exempt us from pursuing a real-world, *ecological* validation of the models [6]. To this aim, we deployed an online Web-based tool^6^ by which clinicians can test the model, by feeding it with clinical values, and considering the sensibleness and usefulness of the indications provided back by the model. After this successful feasibility study, we will conceive proper external validation tasks and undertake an ecological validation to assess the cost-effectiveness and utility of these models for the screening of COVID-19 infection in all the real-world settings (e.g., hospitals, workplaces) where routine blood tests are a viable test of choice.

## Data Availability

Data will be made available after publication in the impacted literature on the Zenodo Platform.

## Declarations

### Ethics approval

Research involving human subjects complied with all relevant national regulations, institutional policies and is in accordance with the tenets of the Helsinki Declaration (as revised in 2013), and was approved by the authors’ Institutional Review Board on the 20th of April.

### Consent to participate and publication

Individuals signed an informed consent authorizing the use of their anonymously collected data for retrospective observational studies (article 9.2.j; EU general data protection regulation 2016/679 [GDPR]), according to the IRCCS San Raffaele Hospital policy (IOG075/2016).

### Availability of data and material

The developed web tool is available at the following address: https://covid19-blood-ml.herokuapp.com/ The complete dataset will be made available on the Zenodo platform as soon as the work gets accepted for publication.

### Code availability

The complete code will be made available on the Zenodo platform as soon as the work gets accepted for publication.

## Footnotes

1 This tool is available at https://covid19-blood-ml.herokuapp.com/

2 A qualitative estimation of the cost of the exams used for this study is 15 euros per test, approximately five times cheaper than rt-PCR testing.

3 IRCCS is the Italian acronym for Scientific Institute for Research, Hospitalization and Healthcare

4 We recall that balanced accuracy is defined as the average of sensitivity and specificity. If accuracy and balanced accuracy significantly differ, the data could be interpreted as unbalanced with respect to class prevalence.

5 We recall here that PPV represents the probability that subjects with a positive screening test truly have the disease.

6 The tool is available at the following address: https://covid19-blood-ml.herokuapp.com/.

